# A Unified Platform for Radiology Report Generation and Clinician-Centered AI Evaluation

**DOI:** 10.1101/2025.07.07.25331018

**Authors:** Zhuoqi Ma, Xinye Yang, Zach Atalay, Andrew Yang, Scott Collins, Harrison Bai, Michael Bernstein, Grayson Baird, Zhicheng Jiao

## Abstract

Generative AI models have demonstrated strong potential in radiology report generation, but their clinical adoption depends on physician trust. In this study, we conducted a radiology-focused Turing test to evaluate how well attendings and residents distinguish AI-generated reports from those written by radiologists, and how their confidence and decision time reflect trust. we developed an integrated web-based platform comprising two core modules: Report Generation and Report Evaluation. Using the web-based platform, eight participants evaluated 48 anonymized X-ray cases, each paired with two reports from three comparison groups: radiologist vs. AI model 1, radiologist vs. AI model 2, and AI model 1 vs. AI model 2. Participants selected the AI-generated report, rated their confidence, and indicated report preference. Attendings outperformed residents in identifying AI-generated reports (49.9% vs. 41.1%) and exhibited longer decision times, suggesting more deliberate judgment. Both groups took more time when both reports were AI-generated. Our findings highlight the role of clinical experience in AI acceptance and the need for design strategies that foster trust in clinical applications. The project page of the evaluation platform is available at: https://zachatalay89.github.io/Labsite.

## 1 Background

In recent years, the rapid advancement of generative AI technologies has significantly propelled the development of the medical and healthcare domains. Among them, large language models have demonstrated remarkable capabilities in language generation and question answering across various tasks, particularly in radiology report generation for X-rays, CT scans, and even MRIs[1]. Studies have shown that generative AI models can achieve notable performance, even reaching accuracy and clinical value comparable to those of radiologists [2]. Despite the promising capabilities of generative AI in radiology report generation, successful integration into clinical workflows hinges on physicians’ acceptance and trust [3, 4]. This is primarily due to the high-stakes nature of medical decision-making, where even occasional errors can have serious consequences. Physicians must not only be confident in the accuracy of AI-generated reports, but also trust their consistency, reliability, and alignment with clinical standards.As a result, beyond technical performance, earning the trust of end-users is a critical prerequisite for the widespread adoption of AI-generated reports in real-world clinical settings.

In this study, we conducted a radiology-oriented Turing test to investigate how well physicians and residents distinguish between AI-generated reports and reports generated by radiologists; and how physicians and residents’ knowledge influence their judgement time and confidence.

## 2 Platform Design

To facilitate both the generation and evaluation of AI-generated radiology reports, we developed an integrated web-based platform comprising two core modules: Report Generation and Report Evaluation, as shown in Figure 1.

**Figure 1:**
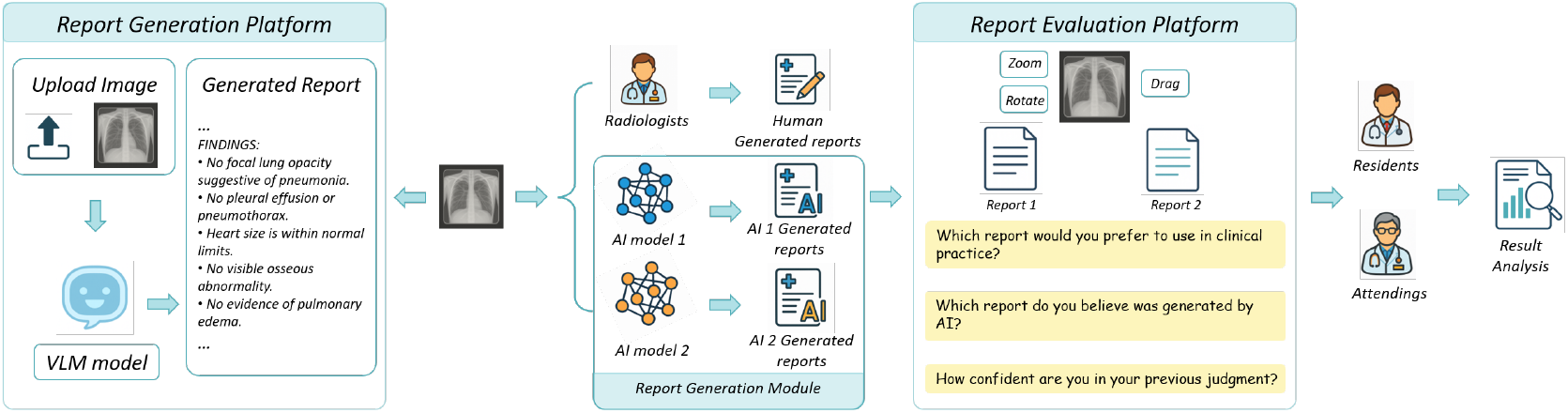
Overview of the radiology report generation and evaluation framework.

### 2.1 Report Generation

This module allows users to upload medical images in various formats (e.g., DICOM, JPG, PNG) via drag-and-drop or file selection. Upon upload, the system automatically processes the image and generates a radiology report using state-of-the-art AI models. The generated report is displayed in a structured format, including metadata (e.g., examination type, technique), findings, and impression. Users can directly edit, copy, or reset the generated report within the interface, enabling efficient post-editing and clinical review.

### 2.2 Report Evaluation

To assess the quality and credibility of AI-generated reports, we designed a Turing test-style evaluation module. Users are presented with two radiology reports side by side, along with the corresponding medical image and an interactive image viewer that supports pan, window/level adjustment, and rotation. Participants are asked to identify which report (if any) they believe is AI-generated, followed by a confidence rating. This enables both qualitative and quantitative assessment of report realism and trustworthiness across different user groups.

By integrating generation and evaluation functionalities within a unified interface, our platform provides a complete end-to-end pipeline for studying human perception of AI-generated medical content, while also supporting clinical usability testing and iterative model improvement.

## 3 Evaluation Pipeline

We use the web-based radiology report evaluation platform to assess physicians’ trust and acceptance of AI-generated radiology reports through a Turing-test-inspired interface. In report evaluation platform, the interface presents a medical image (e.g., an X-ray or CT slice) at the top, accompanied by two diagnostic reports labeled Report 1 and Report 2. Each report contains detailed findings describing various anatomical and pathological observations. The physician is asked to review both reports and the corresponding image, then respond to three key questions: (1) Preference – Which report would you prefer to use in clinical practice? (2) AI Identification – Which report do you believe was generated by AI? (3) Confidence – How confident are you in your previous judgment? To ensure fair evaluation, report identities (AI vs. human) are randomly shuffled across cases. Also, report formatting is standardized to reduce stylistic cues. No model or author identifiers are exposed during the task.

48 X-ray cases and radiologists’ reports were retrieved from Rhode Island Hospital. All data were collected anonymously to ensure participant privacy. For each X-ray case, we generate AI reports using two state-of-the-art methods[5, 6]. The cases were evenly divided into three comparison groups based on report generation methods: radiologist vs. AI model 1, radiologist vs. AI model 2, and AI model 1 vs. AI model 2 (16 each condition).

Eight participants (4 attendings and 4 residents) participated in this experiment. After reading the provided case, participants were asked to identify which report was AI-generated. Additionally, they provided Likert scale evaluations on a scale of 1(low confidence) to 5(high confidence) on their confidence in these judgements. Finally, participants indicate which report they would be more inclined to adopt in clinical practice. This study was IRB approved.

## 4 Results

In Table 1, we report the accuracy, time and confidence between residents and attendings. Attendings were correct 49.9% and residents 41.1% of the time in identifying AI-generated reports, both higher than chance (33.3%), indicating that radiologists largely beat the Turing test and more so than residents. Attendings were more likely to distinguish AI-generated reports from radiologist-generated reports than residents, and both groups were able to distinguish over chance. This suggests that clinical experience helps mitigate bias toward AI-generated content, whereas residents are more prone to perception-driven preference shifts. Attendings exhibited longer average response times (56.84 seconds) compared to residents (31.86 seconds), suggesting a more analytical decision-making approach, p<.01. Furthermore, both groups demonstrated increased response times (attendings by 18.87% and residents by 29%) when evaluating cases in which both reports were AI-generated, indicating that the presence of fully synthetic content introduces greater cognitive load and uncertainty, prompting more prolonged evaluation. Attendings showed slightly more confidence than residents, but this was not significant. For both groups, for every one-unit increase in confidence, the odds of being correct increased 20%, p=.025.

**Table 1:** Performance Comparison Between Residents and Attendings

| Test Case Type | Participants | Accuracy | Mean Response Time (s) | Confidence (Correct) | Confidence (Incorrect) |
| --- | --- | --- | --- | --- | --- |
| Total | Residents | 41.15% | 31.86 | 3.67 | 3.59 |
| Total | Attendings | 48.96% | 56.84 | 3.90 | 3.65 |
| “Both are AI” | Residents | 32.81% | 37.50 | 3.19 | 3.76 |
| “Both are AI” | Attendings | 40.63% | 63.57 | 3.58 | 3.79 |
| “One is AI” | Residents | 45.31% | 29.05 | 3.84 | 3.49 |
| “One is AI” | Attendings | 53.13% | 53.48 | 4.03 | 3.57 |

## 5 Conclusion

Our findings reveal important differences between attendings and residents in terms of acceptance and bias toward AI-generated reports, highlighting the need for targeted strategies to foster trust in AI tools and ensure their safe and effective integration into clinical workflows.

## Data Availability

All data produced in the present study are available upon reasonable request to the authors

https://zachatalay89.github.io/Labsite/

## References

[1] Cheng-Yi Li, Kao-Jung Chang, Cheng-Fu Yang, Hsin-Yu Wu, Wenting Chen, Hritik Bansal, Ling Chen, Yi-Ping Yang, Yu-Chun Chen, Shih-Pin Chen, et al. Towards a holistic framework for multimodal llm in 3d brain ct radiology report generation. Nature Communications, 16(1):2258, 2025.

[2] Eun Kyoung Hong, Byungseok Roh, Beomhee Park, Jae-Bock Jo, Woong Bae, Jai Soung Park, and Dong-Wook Sung. Value of using a generative ai model in chest radiography reporting: a reader study. Radiology, 314(3):e241646, 2025.

[3] Ryutaro Tanno, David GT Barrett, Andrew Sellergren, Sumedh Ghaisas, Sumanth Dathathri, Abigail See, Johannes Welbl, Charles Lau, Tao Tu, Shekoofeh Azizi, et al. Collaboration between clinicians and vision–language models in radiology report generation. Nature Medicine, 31(2):599–608, 2025.

[4] Shruthi Shekar, Pat Pataranutaporn, Chethan Sarabu, Guillermo A Cecchi, and Pattie Maes. People overtrust ai-generated medical advice despite low accuracy. NEJM AI, page AIoa2300015, 2025.

[5] Kang Liu, Zhuoqi Ma, Xiaolu Kang, Zhusi Zhong, Zhicheng Jiao, Grayson Baird, Harrison Bai, and Qiguang Miao. Structural entities extraction and patient indications incorporation for chest x-ray report generation. In International Conference on Medical Image Computing and Computer-Assisted Intervention, pages 433–443. Springer, 2024.

[6] Kang Liu, Zhuoqi Ma, Xiaolu Kang, Yunan Li, Kun Xie, Zhicheng Jiao, and Qiguang Miao. Enhanced contrastive learning with multi-view longitudinal data for chest x-ray report generation. arXiv preprint arXiv:2502.20056, 2025.

